# Circulating white blood cell traits and colorectal cancer risk: A Mendelian randomization study

**DOI:** 10.1101/2023.03.03.23286764

**Authors:** Andrei-Emil Constantinescu, Caroline J Bull, Nicholas Jones, Ruth Mitchell, Kimberley Burrows, Niki Dimou, Stéphane Bézieau, Hermann Brenner, Daniel D Buchanan, Mauro D’Amato, Mark A Jenkins, Victor Moreno, Rish K Pai, Caroline Y Um, Emily White, Neil Murphy, Marc Gunter, Nicholas J Timpson, Jeroen R Huyghe, Emma E Vincent

## Abstract

Observational studies have suggested a protective role for eosinophils in colorectal cancer (CRC) development and implicated neutrophils, but the causal relationships remain unclear. Here, we aimed to estimate the causal effect of circulating white blood cell (WBC) counts (N = ∼550,000) for basophils, eosinophils, monocytes, lymphocytes and neutrophils on CRC risk (N = 52,775 cases and 45,940 controls) using Mendelian randomization (MR). For comparison, we also examined this relationship using individual-level data from UK Biobank (4,043 incident CRC cases and 332,773 controls) in a longitudinal cohort analysis. The inverse-variance weighted (IVW) MR analysis suggested a protective effect of increased basophil count and eosinophil count on CRC risk [OR per 1-SD increase: 0.88, CI(95%): 0.78-0.99, *P*=0.04; OR: 0.93, CI(95%): 0.88-0.98, *P*=0.01]. The protective effect of eosinophils remained [OR per 1-SD increase: 0.88, CI(95%): 0.80-0.97, *P*=0.01] following adjustments for all other WBC subtypes, to account for genetic correlation between the traits, using multivariable MR. A protective effect of increased lymphocyte count on CRC risk was also found [OR: 0.84, CI(95%): 0.76-0.93, *P*=6.70e-4] following adjustment. Consistent with MR results, a protective effect for eosinophils in the cohort analysis in the fully adjusted model [RR per 1-SD increase: 0.96, CI(95%): 0.93-0.99, *P*=0.02] and following adjustment for the other WBC subtypes [RR: 0.96, CI(95%): 0.93-0.99, *P*=0.001] was observed. Our study implicates peripheral blood immune cells, in particular eosinophils and lymphocytes, in CRC development, highlighting a need for mechanistic studies to interrogate these relationships.

**What is already known of this topic:** While previous observational studies have suggested a protective role for eosinophils in colorectal cancer development and implicated neutrophils, whether changes in the levels of circulating white blood cells causes colorectal cancer has not been explored.

**What this study adds:** Our study is the first to use Mendelian randomization (MR) to investigate this relationship. In parallel, for comparison, we also conduct the largest cohort study to date on the topic. We found evidence to suggest that elevated eosinophil and lymphocyte count may have a protective effect on CRC risk, adding new insights into the pathogenesis of CRC.

**How this study might affect research, practice or policy:** Our findings will encourage further mechanistic exploration to understand the biological mechanisms underpinning our findings, which may lead to new therapeutic approaches or risk reduction strategies.

## Introduction

Colorectal cancer (CRC) accounts for over 10% of all worldwide cancer cases and is the second leading cause of cancer-related deaths globally [1–3]. Overall, the number of CRC cases is rising [4] and is alarmingly on the rise in younger people (aged <50 years) [5–7]. Given current challenges, and the estimation that 50% of CRC cases may be preventable [8], a focus on identifying novel risk factors, and subsequent prophylactic and treatment options is warranted to limit the future healthcare burden.

White blood cells (WBCs) are commonly measured in routine blood tests and are divided into five subtypes: basophils, eosinophils, lymphocytes, monocytes and neutrophils [9]. Alterations to circulating WBC counts, have been found to play a role in disease risk, severity, and progression, CRC development and mortality [10–15]. For example, higher circulating counts of basophils and eosinophils, cells with a role in IgE-mediated immunity, have been associated with reduced CRC risk and increased survival [16–20]. Higher absolute lymphocyte count [such as T, B and natural killer (NK) cells] has also been associated with better overall survival of CRC [21,22]. By contrast, a high absolute monocyte count was found to be associated with worse CRC survival [21,22], consistent with their potential role in tumour progression and metastasis [23]. Finally, neutrophils, a critical part of the innate immune system [24], have also been associated with poor CRC overall survival [21,22].

Traditional (i.e. cross-sectional, case-control or cohort) studies account for the majority of epidemiologic analyses undertaken on CRC, which can suffer from certain limitations, such as confounding and reverse causation which can bias effect estimates [25,26]. Mendelian randomization (MR) is a method in genetic epidemiology which could overcome these issues by using single-nucleotide polymorphisms (SNPs) to proxy for an exposure of interest to estimate the effect of an exposure on an outcome [27,28]. It operates akin to a randomized control trial (RCT), as alleles are randomly assigned at birth [29]. We have used MR to estimate the total effect of each WBC trait on CRC [30], and, to account for corelation between WBC traits, multivariable (MV) MR to estimate the direct effect of these exposures on CRC by adjusting for their shared genetic architecture [30].

In this study we aimed to investigate the relationship between circulating WBC subtypes and CRC incidence. Firstly, an MR analysis was undertaken using the most comprehensive genome-wide association study (GWAS) for both WBC subtype counts and incident CRC available. Secondly, for comparison, we ran the largest longitudinal cohort study between WBCs and CRC to date using UK Biobank. Together these analyses allow us to compare between genetically proxied and observational estimates to thoroughly explore the relationship between circulating WBC counts and CRC development.

## Methods

### Study design

We aimed to investigate the relationship between circulating WBCs and CRC using a genetic epidemiologic and complementary observational approach. First, MR analyses were undertaken to estimate the effect of WBC subtype counts on CRC. We then performed a multivariable MR (MVMR) analysis where the direct effect of each WBC subtype count was estimated by adding all five WBC subtypes into the model (**Figure 1A**). STROBE-MR guidelines were followed (**STROBE-MR Supplement**) [31]. Here, units were interpreted as odds ratio (OR) for CRC per a normalized standard deviation (1-SD) increase in WBC count. Next, a prospective longitudinal cohort study was undertaken using UK Biobank individual-level data. Here, subtype specific WBC counts were studied individually, and then each was adjusted for each of the other traits (**Figure 1B**). STROBE guidelines were followed (**STROBE Supplement**). Here, units were interpreted as risk ratio (RR) for CRC per a normalized standard deviation (1-SD) increase in WBC count.

**Figure 1.**
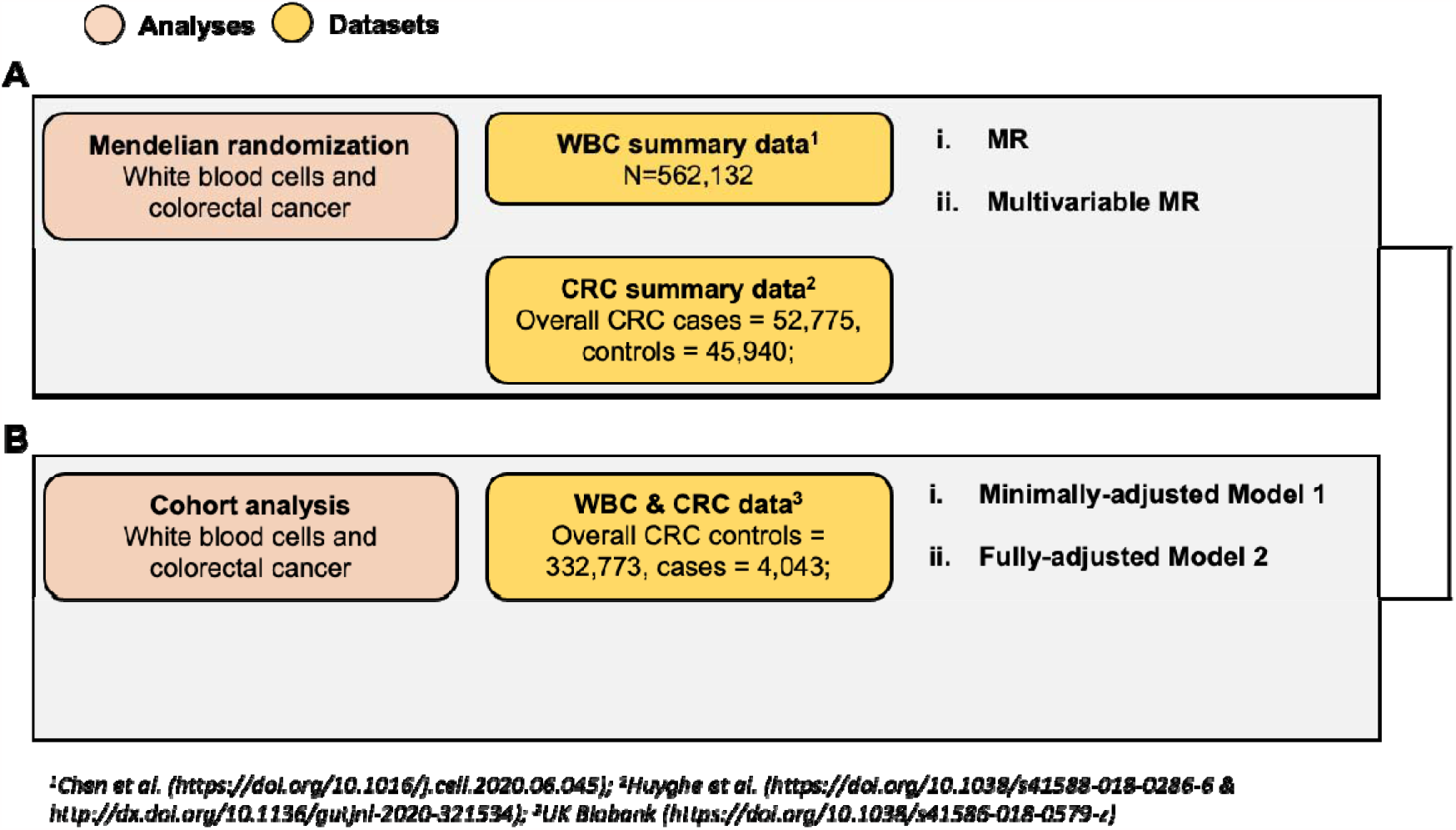
Study design. We triangulated findings from two study designs: a Mendelian randomization analysis(A) and a longitudinal cohort analysis (B) to estimate the casual effect of WBC on CRC risk.

### WBC count GWAS data

Summary statistics for WBC subtype counts were obtained from the “Blood Cell Consortium” (BCX) meta-analysis, the largest study being UKBB (N=∼562,243) [32]. Genetic sex, age, age^2^, study-specific covariates and PCs 1 to 10 were used as covariates. A brief description of each study included in the meta-analysis is available in **Supplementary Table 1**. Only variants which did not display heterogenous effect across studies were selected. Specific details on QC steps and association testing are available in the source manuscripts. Summary statistics for WBC counts were downloaded from: http://www.mhi-humangenetics.org/en/resources/.

### CRC GWAS data

GWAS summary statistics for overall CRC risk and by anatomical subsite were taken from the most comprehensive meta-analyses to date [33]: colon cancer (split into proximal and distal colon cancer) and rectal cancer. Sixty-six studies, which include those of the Genetics and Epidemiology of Colorectal Cancer Consortium (GECCO), Colorectal Cancer Transdisciplinary Study (CORECT), and Colon Cancer Family Registry (CCFR) consortia were meta-analysed [34–36]. The final sample was predominantly of European ancestry with 5.36% of East Asian ancestry. These were included due to their similar genetic architecture with regards to CRC risk [37]. Genetic sex, age, study-specific variables and PCs were used as covariates. An overview of all consortia included in the CRC meta-analysis is available in **Supplementary Table 2**, and a breakdown of the sample sizes for each set of CRC summary statistics is presented in **Supplementary Table 3**.

### Genetic data processing

To select valid MR instruments, summary statistics for exposures were processed using the “TwoSampleMR” R package [38,39]. The presence of correlated instruments, i.e. those in linkage disequilibrium (LD), can bias MR estimates [40]. Therefore, the exposure SNPs were clumped (r^2^=0.001, window=10Mb, P-value threshold=5e-8) using the 1000 Genomes European dataset [41] as a reference panel. Following this step, the exposure and outcome datasets were “harmonised” i.e. had their effect alleles placed on the same reference strand [42]. SNPs with incorrect but unambiguous strand references were corrected, while those with ambiguous strand references were removed. Similarly, palindromic SNPs with inferable strands were kept.

### MR analysis of WBC counts on CRC risk

Primary MR analyses were undertaken using the inverse-variance weighted (IVW) method, which is the fixed-effects meta-analysis of the estimated effect of all exposure SNPs on the CRC outcome [43]. Conditional F-statistics were calculated to detect weak instrument bias [44] for each exposure SNP as previously described [45]. Several sensitivity analyses were undertaken to compare with the main IVW estimates. The presence of vertical pleiotropy i.e. when a trait is downstream of the genetic variant but on the same biological pathway as the exposure [46], was measured using Cochran’s Q heterogeneity test [47]. Horizontal pleiotropy, when some or all instruments for a trait acts through a different pathway to the exposure [46], can violate one of the main MR assumptions. A number of sensitivity MR analyses were undertaken to identify horizontal pleiotropy: MR-Egger (where the regression intercept is not constrained to zero) [48], Weighted median (the median of all SNP ratio estimates, where each ratio is weighted by the inverse of the variance) [49], Weighted mode (assumes that the most frequent estimate in a set of instruments is zero) [50] and MR-PRESSO (detects individual SNPs contributing to horizontal pleiotropy) [51]. The direction of the causal relationship between WBC traits and CRC risk was tested using the MR Steiger method, which uses Steiger’s test to test the difference between the Pearson correlations of genetic variants with both the exposure and outcome [39].

### Multivariable MR analysis of WBC counts on CRC risk

The IVW method was used for the MVMR analysis. First, a pair-wise analysis between all five WBC subtype counts was undertaken, where the proportion of variance explained (PVE) for SNPs used to instrument a WBC trait was estimated in the other four WBC subtypes using previously described methodology [45]. The direct effect of each WBC subtype was estimated by adding in all five WBC subtypes into the MVMR model. Bias arising from weak instruments was also determined here. This was undertaken using methodology described by Sanderson et al., where a generalized version of Cochran’s Q was employed to evaluate instrument strength [52]. Standard Cochran’s Q statistic [47] was calculated to detect the presence of heterogeneity. For those traits with an F-statistic <10, a follow-up MVMR analysis was done accounting for the presence of weak instruments.

### UK Biobank phenotypic data

Between the years of 2007 and 2010, UKBB participants visited assessment centres (N=22) throughout the UK [53,54]. The individuals had their health records linked, their genomes sequenced, and underwent multiple evaluations, such as self-report questionnaires and medical examinations [53,54]. The latter includes the analysis of blood cell samples using Beckman Coulter LH750 instruments designed for high throughput screening [55]. Total WBC count and WBC subtype percentage (%) were measured, with absolute WBC subtype count derived as “WBC subtype % / 100 x total WBC” and expressed as 10^9^ cells/Litre [55]. The blood sampling date variable was split into year, month, day, and minutes (passed since the start of the day of the appointment visit). Additional variables were gathered including recruitment centre, sampling device ID, age, genetic sex, principal components 1 to 10, BMI, Townsend deprivation index, smoking and alcohol drinker status (self-report questionnaire – UKBB codes 20116 and 20117). CRC cases were identified through hospital inpatient records coded to the 10^th^ version of the International Classification of Disease (ICD-10).

### Filtering and selection criteria

The UKBB dataset underwent a series of steps prior to further analyses. Withdrawn participants and those of non-European ancestry were excluded. Viable controls and incident CRC cases were defined using methodology previously described by Burrows et al. [56] (**Supplementary Table 4**). Here, we defined incident CRC cases as those diagnosed at least one year after blood sampling. Participants with no WBC measurement data or sampling date were removed, as were those who were known to be pregnant, have chronic conditions (e.g. HIV, blood cancers, thalassaemia), or undergoing erythropoietin treatment, as in Astle et al. [57] and Chen et al. [32], given the effects of these traits on WBC measurements. Those with acute conditions (e.g. upper respiratory infections) diagnosed less than 3 months prior to blood sampling were excluded. Finally, missing values in “Townsend Deprivation Index”, “Body mass index”, “Smoking status” and “Alcohol drinker status” variables were removed.

### Cohort study between WBC count and CRC

We conducted a cohort analysis between circulating WBCs and incident CRC. WBC count values were log-transformed, after which they were adjusted for the following covariates: assessment centre, sex, age, age^2^, PCs 1 to 10, as in the Chen et al. GWAS [32]. The resulting residuals were rank-inverse normal transformed and then used in a logistic regression on CRC incidence. This main observational analysis was termed “Model 1”, a minimally adjusted model. An additional fully adjusted analysis was undertaken, termed “Model 2”, where BMI, Townsend DI, smoker status and alcohol drinker status were added as additional covariates. Following this, another pair of analyses was run, where all five WBC subtype counts were added together into the model to reduce potential bias due to their correlated values. Analyses where each WBC trait was studied individually were termed as “univariable”, while those where they were added together were termed as “multivariable”.

### Working environment

All analyses were performed with R version 4.1.2 (Bird Hippie) in a Linux environment supported by the University of Bristol’s Advanced Computing Research Centre (ACRC). Genetic data preparation, as well as the MR analyses, were undertaken with the “TwoSampleMR” R package [38,39]. The MVMR analyses were undertaken with the “MVMR” R package [58]. Scripts associated with this study are available on GitHub: https://github.com/andrewcon/wbc-crc.

## Results

### Effect of WBC count on CRC

Before performing the MR analysis, the average F-statistic for each WBC trait was estimated to detect the presence of weak instrument bias, which is generally indicated by an average F-statistic < 10 [44]. For overall CRC, these were 64.48 (basophil count, 171 SNPs), 124.72 (eosinophil count, 396 SNPs), 105.85 (lymphocyte count, 444 SNPs), 147.44 (monocyte count, 477 SNPs), 98.84 (neutrophil count, 387 SNPs), indicating strong MR instruments (**Supplementary Table 5**).

The IVW method showed evidence of a protective effect of basophil count (1-SD increase in WBC count OR: 0.88, 95% CI: 0.78-0.99, P-value: 0.037) for CRC risk (**Figure 2, Supplementary Figure 1**). For eosinophil count, the IVW method showed evidence of a protective effect for CRC risk (OR: 0.93, 95% CI: 0.88-0.98, P-value: 0.012) (**Figure 2, Supplementary Figure 1**). These results were confirmed by the MR-Egger, weighted median and MR-PRESSO sensitivity methods (**Supplementary Table 6**). The IVW method also showed a protective effect of eosinophil count for CRC risk (OR: 0.91, 95% CI: 0.85-0.99, P-value: 0.021) in females. All results, including those for sex- and site-specific CRC, are available in **Supplementary Table 6**.

**Figure 2.**
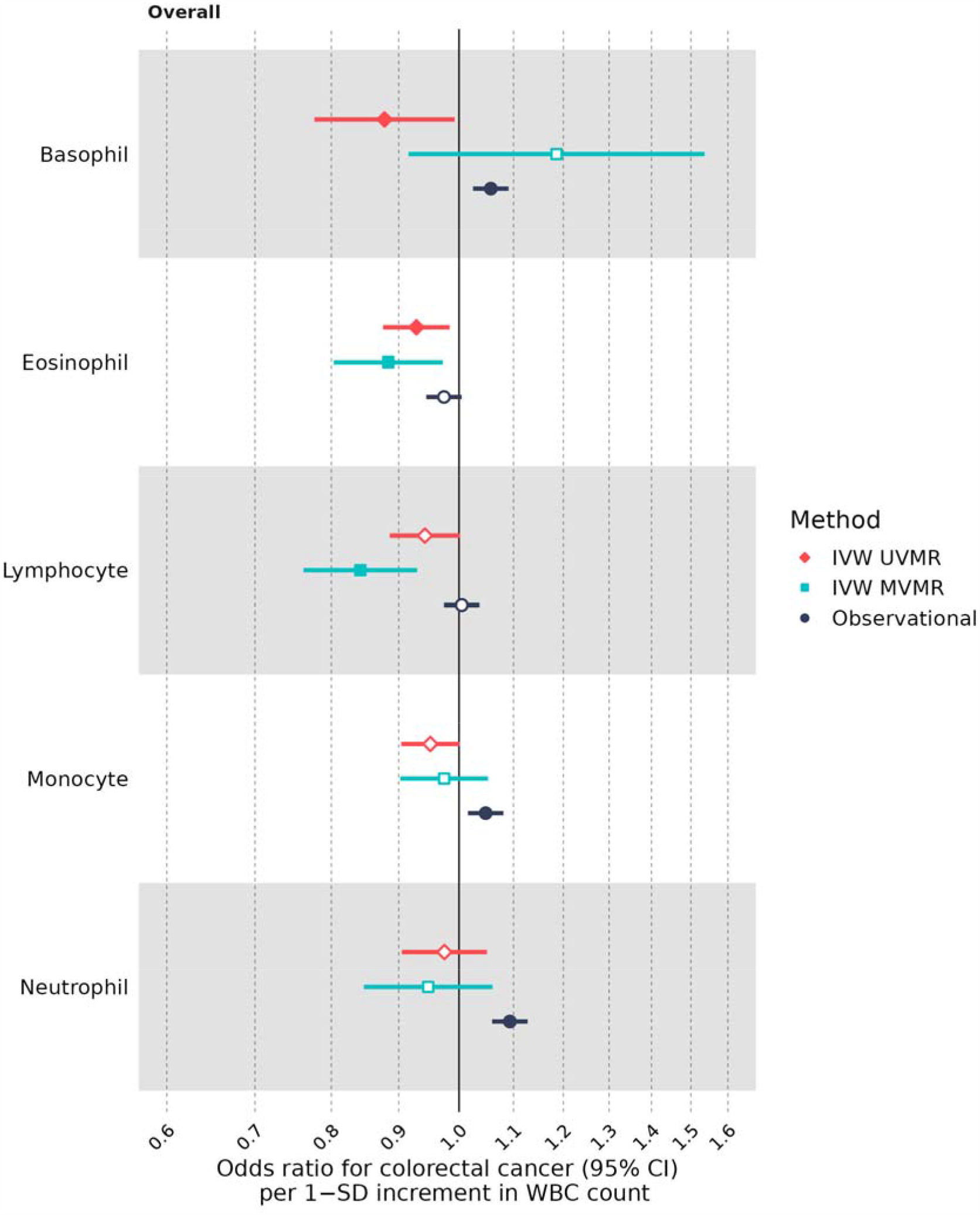
The relationship between WBC count and CRC risk based on UV, MV two-sample MR and cohort observational analyses. Each WBC trait is presented on the X-axis. The estimated effect is presented on the Y-axis. Point estimates were filled where the P-value was less than 0.05. Results are interpreted as ORs (95% CI) for CRC risk per 1-SD normalized increment in WBC count.

In site specific CRC analyses, the univariable IVW method demonstrated evidence for a protective effect of basophil count on colon (OR: 0.85, 95% CI: 0.74-0.98, P-value: 0.022) and distal colon (OR: 0.82, 95% CI: 0.70-0.97, P-value: 0.019) cancer (**Supplementary Table 6**). For eosinophil count, the main IVW analysis provided evidence for a protective effect for colon (OR: 0.90, 95% CI: 0.84-0.96, P-value: 0.001), proximal colon (OR: 0.89, 95% CI: 0.82-0.96, P-value: 0.003) and distal colon (OR: 0.89, 95% CI: 0.82-0.97, P-value: 0.007) cancer (**Supplementary Table 6**). These results were mostly supported by sensitivity analyses. There was a protective effect of total WBC count on colon (OR: 0.91, 95% CI: 0.85-0.99, P-value: 0.02) and proximal colon (OR: 0.90, 95% CI: 0.82-0.98, P-value: 0.015) cancer (**Supplementary Table 6**).

Next, the presence of vertical and horizontal pleiotropy in the MR analyses was analysed. Cochran’s heterogeneity test indicated the presence of heterogeneity in all but one WBC trait-CRC pair (basophil count-male CRC, P_HET_=0.104) (**Supplementary Table 7**). Following this, the MR-Egger test for horizontal pleiotropy was performed. Here, evidence for this type of pleiotropy was identified for eosinophil count and colon (P_PLT_=0.018), distal colon (P_PLT_=0.015), female (P_PLT_=0.049), male (P_PLT_=0.041) and overall (P_PLT_=0.015) CRC risk, suggesting a possible bias of MR estimates. This was also the case for lymphocyte count in female CRC (P_PLT_=0.016) (**Supplementary Table 7**). Although the MR-PRESSO method identified the presence of SNP horizontal-pleiotropic outliers, there was little evidence that the removal of these outliers contributed to a notable shift in the point estimates (**Supplementary Table 8**).

### Multivariable MR of WBC count on CRC

A pair-wise analysis of the proportion of variance explained (PVE) for SNPs instrumenting each WBC trait indicated a low PVE for each of the other WBC traits, with the exception of basophil count (2.44% vs. 2.39% when instrumenting neutrophil count) (**Supplementary Table 9**). The overall results indicated that statistical power should not suffer to a large degree by adding all five WBC subtype counts into the MVMR analysis. Here, the MVMR IVW method estimated a protective effect of eosinophil count (OR: 0.88, 95% CI: 0.80-0.97, P-value: 0.011) and lymphocyte count (OR: 0.84, 95% CI: 0.76-0.93, P-value: 0.0007) on CRC risk (**Figure 2**). Regarding sex-specific estimates, eosinophil count was protective for CRC risk (OR: 0.83, 95% CI: 0.73-0.94, P-value: 0.004) in females, this was also the case for lymphocyte count (OR: 0.76, 95% CI: 0.67-0.87, P-value: 6.46E-05). Eosinophil count was estimated to have a direct protective effect on colon (OR: 0.84, 95% CI: 0.75-0.94, P-value: 0.002), proximal colon (OR: 0.92, 95% CI: 0.85-1.00, P-value: 0.042) and distal colon (OR: 0.88, 95% CI: 0.74-1.00, P-value: 0.049) cancer (**Supplementary Table 10**). Lymphocyte count was estimated to have a direct protective effect on colon (OR: 0.85, 95% CI: 0.76-0.96, P-value: 0.007), distal colon (OR: 0.77, 95% CI: 0.67-0.88, P-value: 0.0001) and rectal (OR: 0.86, 95% CI: 0.75-0.98, P-value: 0.022) cancer anatomical subsites (**Supplementary Table 10**).

Sensitivity analyses were undertaken to assess heterogeneity and the presence of weak instruments in the MVMR analysis. There was evidence of heterogeneity in all WBC trait-CRC pairs (**Supplementary Table 11**). Conditional F-statistics showed evidence of weak instruments (F<10) for basophil count (**Supplementary Table 11**). Based on these results, an additional MVMR analysis was run adjusting for weak instruments for basophil count (overall CRC OR_Weak_: 1.3; male CRC OR_Weak_: 1.2; female CRC OR_Weak_: 1.7; colon cancer OR_Weak_: 1.5; proximal colon cancer OR_Weak_: 1.5; distal colon cancer OR_Weak_: 1.5; rectal cancer OR_Weak_: 1.6) on CRC risk (**Supplementary Table 11**).

### Cohort observational analysis between WBC count and CRC

336,816 UKBB participants remained after passing filtering and selection criteria (Supplementary Figure 2). Of these, 332,773 were controls and 4,043 were incident CRC cases. When split by genetic sex, there were 154,629 male and 178,144 female controls and 2,316 male and 1,727 female cases. Those with CRC were more likely to be male (57% vs. 46%), had a higher average age (60.7 vs. 55.8 years), slightly higher BMI (28.0 vs. 27.4 kg/m^2^) and were more likely to have been cigarette smokers in the past (46% vs. 55% never smokers and 44% vs. 34% pervious smokers) (Table 1).

**Table 1.** Baseline characteristics of UK Biobank study sample.

| **Characteristic** | **Control, N = 332,773<sup>1</sup>** | **Case, N = 4,043<sup>1</sup>** |
| --- | --- | --- |
| Sex |  |  |
| Female | 178,144 / 332,773 (54%) | 1,727 / 4,043 (43%) |
| Male | 154,629 / 332,773 (46%) | 2,316 / 4,043 (57%) |
| Age (years) | 55.8 (8.1) | 60.7 (6.6) |
| Body mass index | 27.4 (4.7) | 28.0 (4.7) |
| Smoking status |  |  |
| Never | 183,987 / 332,773 (55%) | 1,861 / 4,043 (46%) |
| Previous | 114,349 / 332,773 (34%) | 1,790 / 4,043 (44%) |
| Current | 34,437 / 332,773 (10%) | 392 / 4,043 (9.7%) |
| Alcohol drinker status |  |  |
| Never | 10,655 / 332,773 (3.2%) | 122 / 4,043 (3.0%) |
| Previous | 10,982 / 332,773 (3.3%) | 140 / 4,043 (3.5%) |
| Current | 311,136 / 332,773 (93%) | 3,781 / 4,043 (94%) |
| Townsend deprivation index | -1.4 (3.0) | -1.6 (3.0) |
| Basophil count | 0.0 (0.1) | 0.0 (0.1) |
| Eosinophil count | 0.2 (0.1) | 0.2 (0.1) |
| Lymphocyte count | 1.9 (0.6) | 1.9 (0.6) |
| Monocyte count | 0.5 (0.2) | 0.5 (0.2) |
| Neutrophil count | 4.2 (1.4) | 4.3 (1.4) |
| Overall WBC count | 6.9 (1.7) | 7.0 (1.8) |
| 1n / N (%); Mean (SD) |  |  |

As a percentage of the total WBC count based on the median values, basophils accounted for 0.3%, eosinophils 2.11%, lymphocytes 28.16%, monocytes 6.78%, and neutrophils 60.39% (**Supplementary Table 12**). A pair-wise correlation matrix between each WBC subtype showed a high correlation between total WBC and neutrophil count, while for the other subtypes the correlation coefficients were equal or below 0.3 (**Supplementary Figure 3**). Batch variables (e.g. blood sample device and sampling date), Townsend DI, and alcohol drinker status explained some of the variance in WBC count (0% to 0.66%). Depending on the WBC subtype, genetic sex explained between 0.23% to 2.93% of the variance, BMI explained between 0.14% to 2.64%, and smoking status explained between 0.44% to 3.85% (**Supplementary Figure 4**).

In the main analysis (“Model 1” - the minimally adjusted model), basophil count was positively associated with CRC risk (RR: 1.06, 95% CI: 1.02-1.09, P-value: 0.0005), as was monocyte (RR: 1.05, 95% CI: 1.02-1.08, P-value: 0.003) and neutrophil count (RR: 1.09, 95% CI: 1.06-1.13, P-value: 1.94E-08) (**Figure 2, Supplementary Table 13**). In “Model 2”, eosinophil count was negatively associated with CRC risk (overall CRC RR: 0.96, 95% CI: 0.93-0.99, P-value: 0.022) (**Supplementary Table 13**).

Observational associations were re-computed by adding all five WBC subtype counts together. In the minimally adjusted “Model 1”, eosinophil count (RR: 0.96, 95% CI: 0.93-0.99, P-value: 0.009) was associated with lower overall CRC odds, while basophil (RR: 1.04, 95% CI: 1.01-1.08, P-value: 0.008) and neutrophil count (RR: 1.08, 95% CI: 1.05-1.12, P-value: 1.92E-06) were associated with an increase in overall CRC risk (**Supplementary Table 14**). These results largely coincided with those from the fully adjusted “Model 2” analyses (**Supplementary Table 14**). All results, including those for sex-specific CRC, are available in **Supplementary Table 14**.

## Discussion

In this study we aimed to estimate the effects of five circulating WBC subtypes on CRC risk by using a combined genetic epidemiologic and longitudinal cohort framework. Through the aid of MVMR, we were able to assess the independent causal effect of WBC counts by adjusting for their shared genetic architecture. Taken together, the evidence across analyses suggests a potential protective effect of increased circulating eosinophil and lymphocyte count on CRC risk.

Consistent with our study, Prizment et al found that eosinophil count (tertiles Q3 and Q2 vs. Q1) was negatively associated with odds of developing colon, but not of rectal cancer [19]. Similar results have been reported for other cancers; Wong et al. reported a negative trend between increasing eosinophil count quartiles and lung adenocarcinoma odds in a UKBB study [59], while a similar study looking at prostate cancer showed a negative association between eosinophil count quintiles Q3-5, as well as a per 1-SD increase in the trait (HR 0.96 vs. OR 0.93 for CRC in our analysis) [60].

Eosinophils have a well-established role in allergic disease, including asthma and allergic rhinitis [61]. Indeed, MR analyses have also reported a causal effect of eosinophil count on allergic disease [57,62] and a recent systematic review investigating the relationship between allergies and cancer reported evidence for a reduced risk of CRC in those with allergic diseases [63].

Here, our results suggest that the immune response through by eosinophils provides protection against tumour development. Indeed, in several neoplasia, including CRC, eosinophils have been found to play an anti-tumourigenic role and are a source of anti-tumourigenic molecules, such as eosinophil-derived neurotoxin (EDN) [64,65]. Experimental studies have also found a tumour-protective effect of IgE [66]. Increased eosinophil recruitment to the CRC tumour site has also been associated with better survival, even when adjusting for the effects of CD8^+^ T-cells [13], and eosinophil-specific granule secretion of granzyme A has been linked with the killing of CRC cells [67].

In addition to eosinophil count, we also found a protective effect of lymphocyte count on CRC risk. While this was not apparent in the MR analysis, the MVMR estimates indicated lower ORs for CRC across all anatomical subsites (proximal colon cancer trended towards protective) with increased lymphocyte counts. The multivariable fully adjusted “Model 2” in the cohort analysis also indicated that there could be a negative association with CRC risk. It is not surprising that we found a protective effect of higher circulating levels of lymphocytes with CRC odds given their established role in combatting tumour development [68]. Tumour-infiltrating lymphocytes (TILs) like CD8+ T-cells help antagonise tumour growth through direct action and recruitment of other immune cells [68]. High levels of TILs were previously associated with better CRC overall survival and disease-free survival [22,68]. In support of our findings, two observational studies found higher lymphocyte counts compared to cases vs. controls a year to six months prior to CRC diagnosis [69,70]. However, these results could indicate production and recruitment of lymphocytes to the site of pre-cancerous or undetected tumours rather than a causal effect.

## Limitations

There are several limitations to this study. With regards to the cohort analysis, only baseline blood measurements were available. This assumes that WBC counts were constant and did not allow us to establish a relationship between a trend in WBC count and its relationship with CRC odds. Nevertheless, baseline WBC count measurements have previously been shown to be associated with disease risk [59,60,71,72], making their study in relation to disease development a worthwhile endeavour. Also, in the cohort analysis, incident CRC cases were defined as those diagnosed at least one year after blood sampling, in order to not diminish the number of cases to a large degree. However, as CRC develops over a long period, our cohort analysis may have not excluded all participants with undiagnosed CRC.

With regards to the MR analysis, the genetic instruments used here proxied for lifetime variation of WBC count. Therefore, the MR analysis cannot be used to infer how large changes over a short timespan might affect CRC development. Regarding the MVMR method, caution should be applied when investigating traits with very weak instruments, as it cannot reliably adjust for those traits [58]. This was the case for basophil count, as the F-statistic was estimated to be between 4.7 and 4.8 (**Supplementary Table 12**). Therefore, despite pointing to an increased detrimental effect compared to the main MVMR analysis, ORs derived from the weak-MVMR analysis should be interpreted with this in mind.

## Conclusion

In summary, the results generated here provide evidence for a protective causal effect of elevated levels of circulating eosinophil and lymphocyte counts on CRC risk. Going forward, additional research is needed to disentangle the biological mechanisms and pinpoint specific pathways through which eosinophils and lymphocytes might protect against CRC development.

## Availability of data and materials

Summary statistics for WBC counts were downloaded from the following website: http://www.mhi-humangenetics.org/en/resources/. The summary-level GWAS data on outcomes used in this study were made available following an application to the Genetics and Epidemiology of Colorectal Cancer Consortium (GECCO): https://www.fredhutch.org/en/research/divisions/public-health-sciences-division/research/cancer-prevention/genetics-epidemiology-colorectal-cancer-consortium-gecco.html.

## Supporting information

STROBE-MR

STROBE

## Contributions

AC, CJB, JH and EEV conceived the idea for the paper. AC conducted the analysis. All authors contributed to the interpretation of the findings. AC, CJB and EEV wrote the manuscript. All authors critically revised the paper for intellectual content and approved the final version of the manuscript.

## Funding information

AC acknowledges funding from grant MR/N0137941/1 for the GW4 BIOMED MRC DTP, awarded to the Universities of Bath, Bristol, Cardiff and Exeter from the Medical Research Council (MRC)/UKRI. NJT is the PI of the Avon Longitudinal Study of Parents and Children (Medical Research Council & Wellcome Trust 217065/Z/19/Z) and is supported by the University of Bristol NIHR Biomedical Research Centre (BRC-1215-2001). NJT acknowledges funding from the Wellcome Trust (202802/Z/16/Z). EEV, CJB, ND and NJT acknowledge funding by the CRUK Integrative Cancer Epidemiology Programme (C18281/A29019). NJT, EEV and CJB work in a unit funded by the UK Medical Research Council (MC_UU_00011/1 & MC_UU_00011/4) and the University of Bristol. EEV and CJB are supported by Diabetes UK (17/0005587) and the World Cancer Research Fund (WCRF UK), as part of the World Cancer Research Fund International grant program (IIG_2019_2009). JRH acknowledges funding by the National Cancer Institute at the U.S. National Institutes of Health (R21CA230486). This work was also supported by the Elizabeth Blackwell Institute for Health Research, University of Bristol, and the Wellcome Trust Institutional Strategic Support Fund (ISSF 204813/Z/16/Z). The funders of the study had no role in the study design, data collection, data analysis, data interpretation or writing of the report.

## Ethics declaration

All participants provided written informed consent, and each study was approved by the relevant research ethics committee or institutional review board.

## Consent for publication

All authors consented to the publication of this work.

## Competing interests

The authors declare no competing interests.

## Supplementary information

Additional file 1

**Supplementary Figures**.

Additional file 2

**Supplementary Tables**.

Additional file 3

**STROBE-MR checklist**.

Additional file 4

**STROBE Cohort checklist**.

Additional file 5

**Full funding and acknowledgements details**.

## Notes

### Competing Interest Statement

The authors have declared no competing interest.

