## Supplemental Figure for "Circulating white blood cell traits and colorectal cancer risk: A Mendelian randomization study"

### SUPPLEMENTARY FIGURES for “Investigating the effect of circulating immune cell counts on colorectal cancer risk”

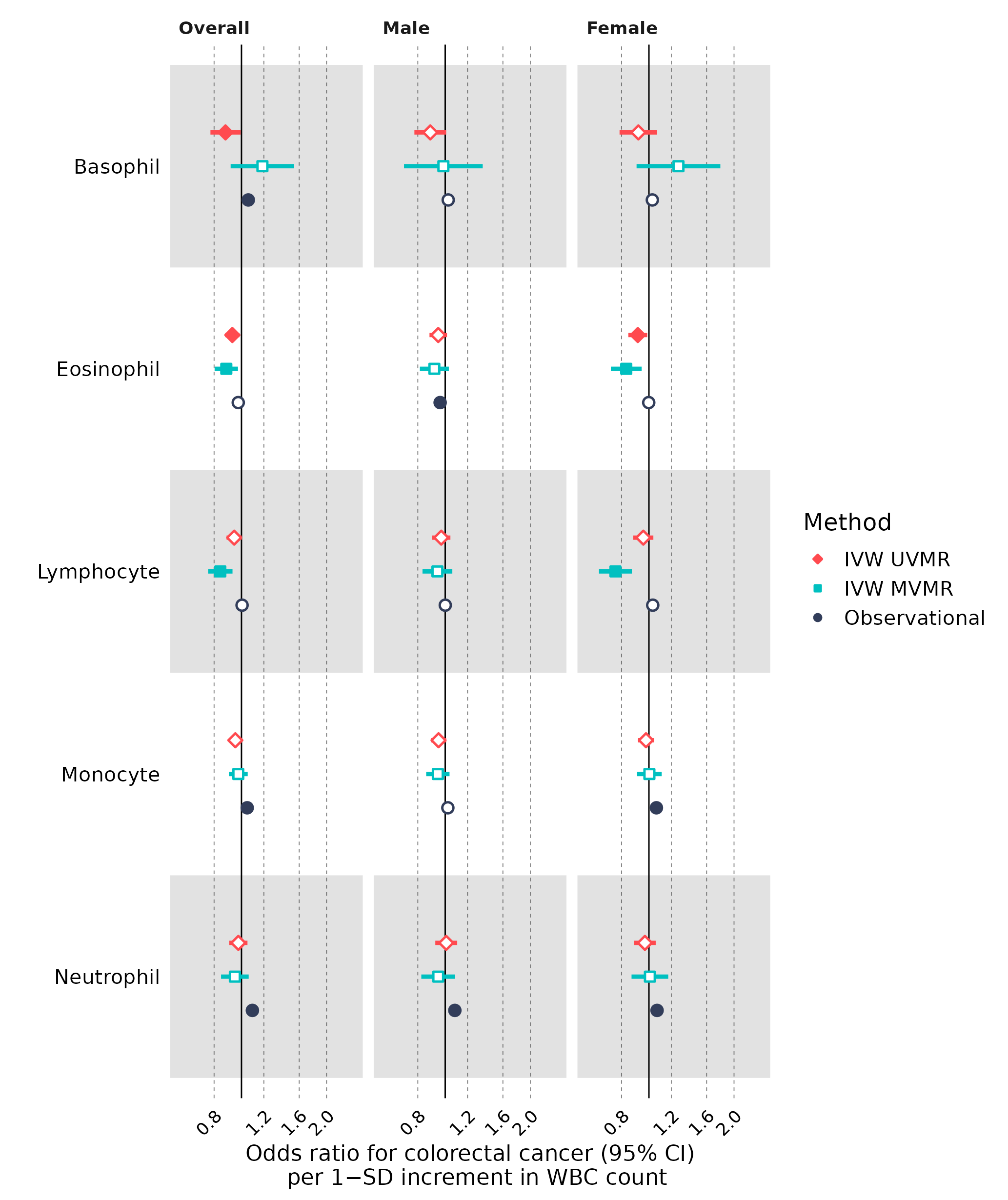

**Supplementary Figure 1. Analyses of WBC count on overall, and by genetic sex CRC.** WBC traits are separated into rows on the X-axis. Each column is an analysis looking at overall, male-specific, and female-specific CRC. The estimated effect given by each method is presented on the Y-axis. Point estimates were filled where the P < 0.05. Results are interpreted as ORs (MR) or RRs (Cohort) (95% CI) for CRC per 1-SD normalized increment in WBC count.

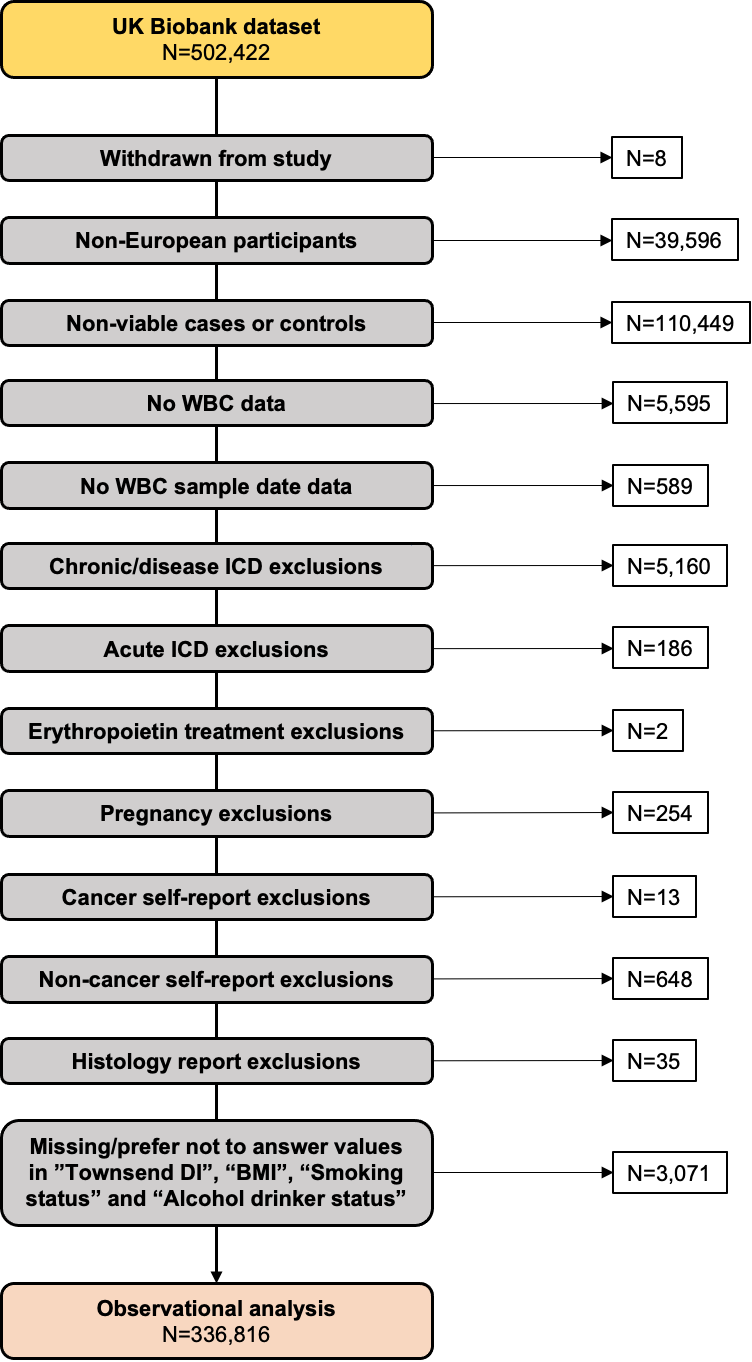

**Supplementary Figure 2. Cohort study sample selection flowchart.**

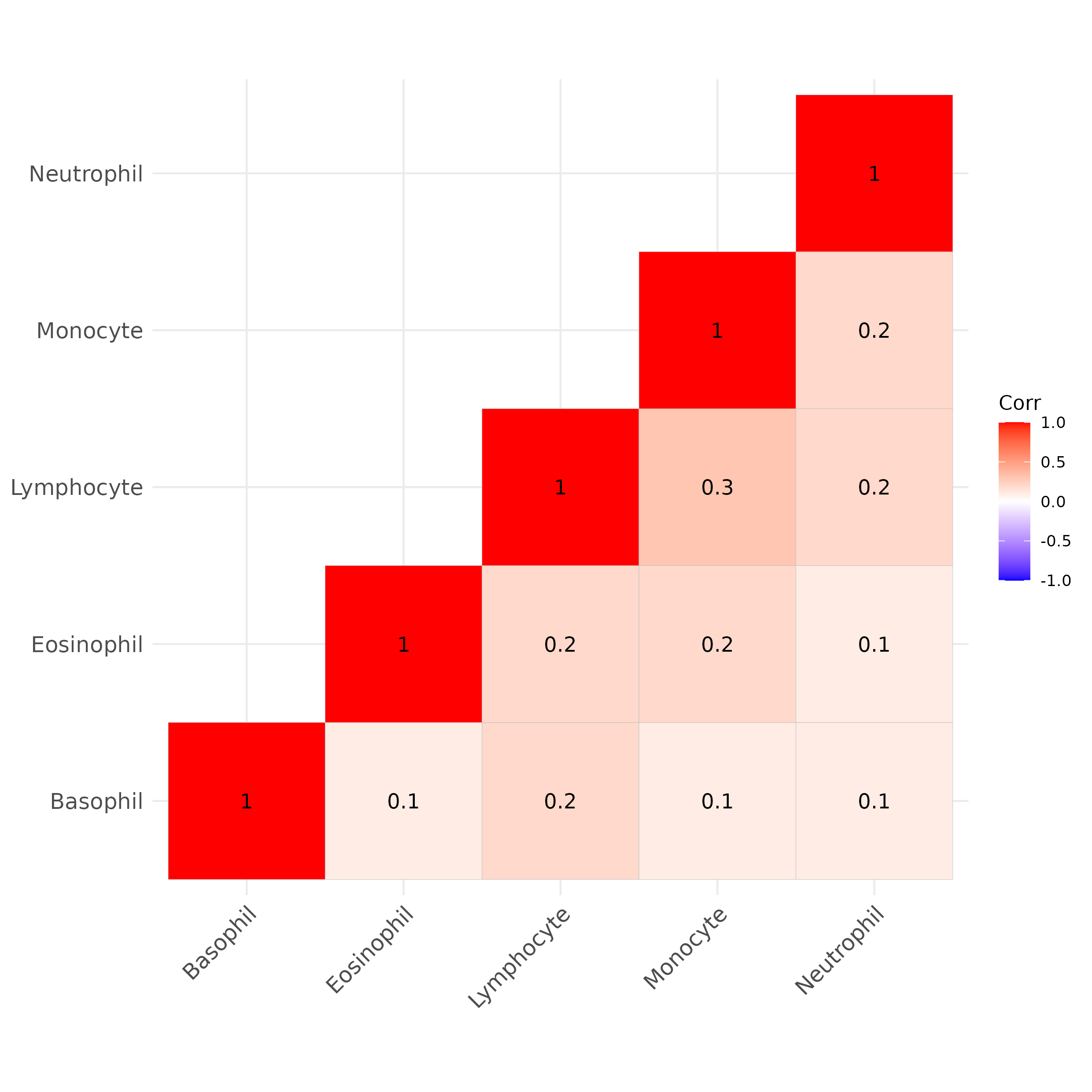

**Supplementary figure 3. Pair-wise correlation matrix.** The total WBC count and WBC subtype counts were analysed between each other using Spearman correlation. WBC traits are on the X and Y axes. The number inside each square represents the correlation coefficient between the studied traits for each WBC trait pair.

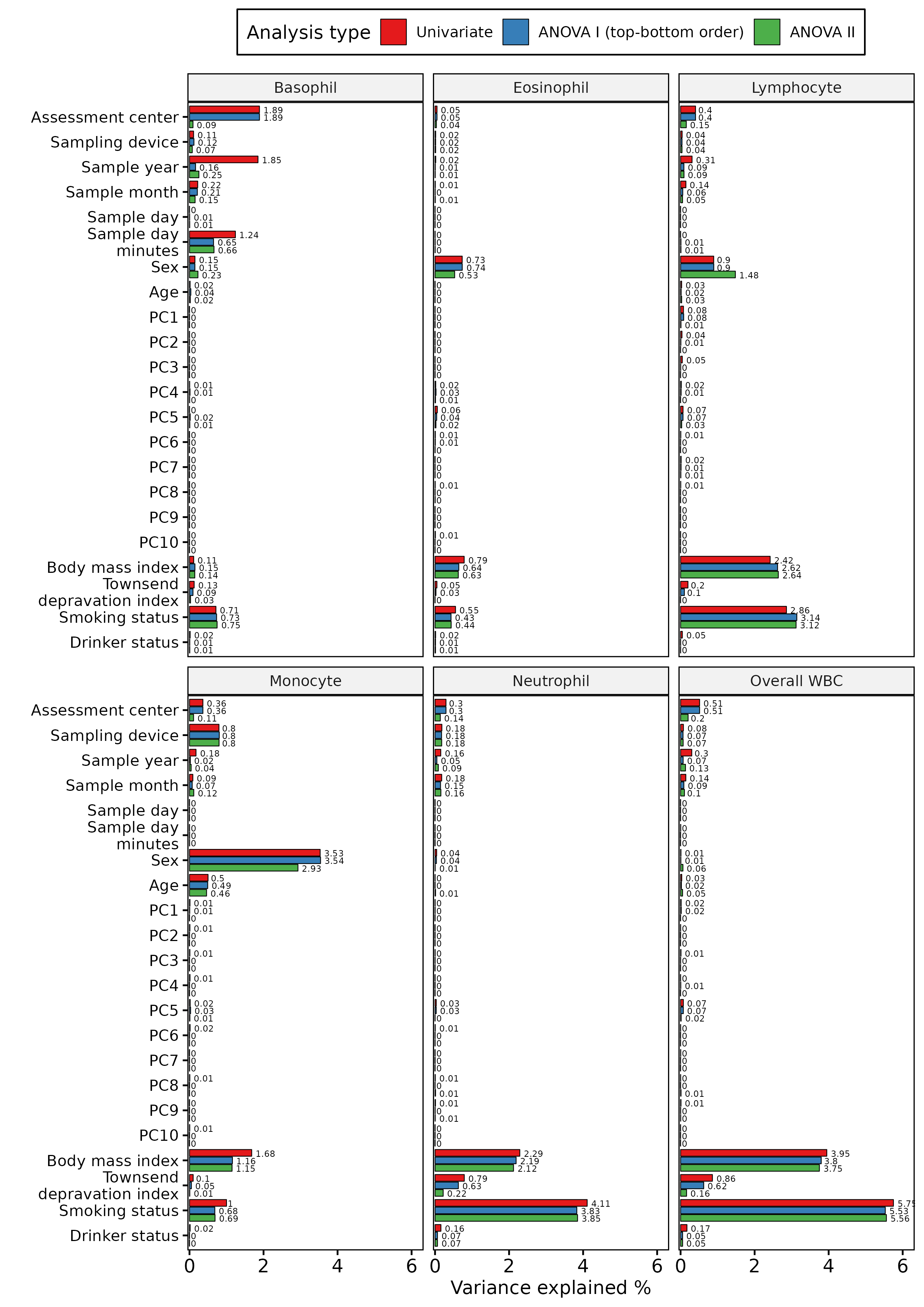

**Supplementary Figure 4. Variance explained by variables on WBC count.** Univariable, type I ANOVA and type II ANOVA were run to determine the variance explained on overall WBC count, as well as each WBC subtype count, which are represented as separate facets in the figure. Studied variables were ordered top-down on the Y-axis, which is the way the type I ANOVA was run. The X-axis represents the variance explained (%) on WBC count for the corresponding variable for each analysis type.
