## Supplementary material for "Circulating white blood cell traits and colorectal cancer risk: A Mendelian randomization study": STROBE-MR

**STROBE-MR checklist of recommended items to address in reports of Mendelian randomization studies**^1^ ^2^

| **Item No.** | **Section** | **Checklist item** | **Page No.** | **Relevant text from manuscript** |
| --- | --- | --- | --- | --- |
| 1 | **TITLE and ABSTRACT** | Indicate Mendelian randomization (MR) as the study’s design in the title and/or the abstract if that is a main purpose of the study | 1-3 |  |
|  | **INTRODUCTION** |  |  |  |
| 2 | **Background** | Explain the scientific background and rationale for the reported study. What is the exposure? Is a potential causal relationship between exposure and outcome plausible? Justify why MR is a helpful method to address the study question | 3-5 |  |
| 3 | **Objectives** | State specific objectives clearly, including pre-specified causal hypotheses (if any). State that MR is a method that, under specific assumptions, intends to estimate causal effects | 6 | Introduction, paragraphs 4,5 |
|  | **METHODS** |  |  |  |
| 4 | **Study design and data sources** | Present key elements of the study design early in the article. Consider including a table listing sources of data for all phases of the study. For each data source contributing to the analysis, describe the following: |  |  |
|  | a) | Setting: Describe the study design and the underlying population, if possible. Describe the setting, locations, and relevant dates, including periods of recruitment, exposure, follow-up, and data collection, when available. | 6-8 | Methods, paragraphs 1-4 |
|  | b) | Participants: Give the eligibility criteria, and the sources and methods of selection of participants. Report the sample size, and whether any power or sample size calculations were carried out prior to the main analysis | 7-8 | Methods, paragraphs 2-4. Supplementary Tables 1-4 |
|  | c) | Describe measurement, quality control and selection of genetic variants | 8 | Methods, paragraphs 2-4. More in the manuscript associated with each study. |
|  | d) | For each exposure, outcome, and other relevant variables, describe methods of assessment and diagnostic criteria for diseases | 7-8 | Methods, paragraphs 2-4. Supplementary Tables 1-4 |
|  | e) | Provide details of ethics committee approval and participant informed consent, if relevant | 22 | Ethics declaration |
| 5 | **Assumptions** | Explicitly state the three core IV assumptions for the main analysis (relevance, independence and exclusion restriction) as well assumptions for any additional or sensitivity analysis | 5 | Introduction, paragraph 4 |
| 6 | **Statistical methods: main analysis** | Describe statistical methods and statistics used | 8-11 | Methods, paragraphs 6,7,11 |
|  | a) | Describe how quantitative variables were handled in the analyses (i.e., scale, units, model) | 6 | Methods, paragraph 1 |
|  | b) | Describe how genetic variants were handled in the analyses and, if applicable, how their weights were selected | 9 | Methods, paragraph 5 |
|  | c) | Describe the MR estimator (e.g. two-stage least squares, Wald ratio) and related statistics. Detail the included covariates and, in case of two-sample MR, whether the same covariate set was used for adjustment in the two samples | 9,10 | Methods, paragraphs 8,9 |
|  | d) | Explain how missing data were addressed | N/A |  |
|  | e) | If applicable, indicate how multiple testing was addressed | N/A |  |
| 7 | **Assessment of assumptions** | Describe any methods or prior knowledge used to assess the assumptions or justify their validity | 9 | Methods, paragraph 7 |
| 8 | **Sensitivity analyses and additional analyses** | Describe any sensitivity analyses or additional analyses performed (e.g. comparison of effect estimates from different approaches, independent replication, bias analytic techniques, validation of instruments, simulations) | 10 | Methods, paragraphs 8,9 |
| 9 | **Software and pre-registration** |  |  |  |
|  | a) | Name statistical software and package(s), including version and settings used | 10 | Methods, paragraph 10 |
|  | b) | State whether the study protocol and details were pre-registered (as well as when and where) | N/A |  |
|  | **RESULTS** |  |  |  |
| 10 | **Descriptive data** |  |  |  |
|  | a) | Report the numbers of individuals at each stage of included studies and reasons for exclusion. Consider use of a flow diagram | 11 | Supplementary Figure 2 |
|  | b) | Report summary statistics for phenotypic exposure(s), outcome(s), and other relevant variables (e.g. means, SDs, proportions) | 11 | Table 1, Supplementary |
|  | c) | If the data sources include meta-analyses of previous studies, provide the assessments of heterogeneity across these studies | 8 | Methods, paragraph 5 |
|  | d) | For two-sample MR:  i.  Provide justification of the similarity of the genetic variant-exposure associations between the exposure and outcome samples  ii.  Provide information on the number of individuals who overlap between the exposure and outcome studies | N/A | 0% overlap between samples |
| 11 | **Main results** |  |  |  |
|  | a) | Report the associations between genetic variant and exposure, and between genetic variant and outcome, preferably on an interpretable scale | N/A | Summary statistics for CRC are not currently publicly available. |
|  | b) | Report MR estimates of the relationship between exposure and outcome, and the measures of uncertainty from the MR analysis, on an interpretable scale, such as odds ratio or relative risk per SD difference | 13-16 | Results, paragraphs 3,4,6,7,10,11. Supplementary Tables 10,14 |
|  | c) | If relevant, consider translating estimates of relative risk into absolute risk for a meaningful time period | N/A |  |
|  | d) | Consider plots to visualize results (e.g. forest plot, scatterplot of associations between genetic variants and outcome versus between genetic variants and exposure) |  | Figure 2,3 |
| 12 | **Assessment of assumptions** |  |  |  |
|  | a) | Report the assessment of the validity of the assumptions | 14,16 | Results, paragraphs 8,12 |
|  | b) | Report any additional statistics (e.g., assessments of heterogeneity across genetic variants, such as *I^2^*, Q statistic or E-value) | 14,16 | Results, paragraphs 8,12 |
| 13 | **Sensitivity analyses and additional analyses** |  |  |  |
|  | a) | Report any sensitivity analyses to assess the robustness of the main results to violations of the assumptions | 14,16 | Results, paragraphs 8,12 |
|  | b) | Report results from other sensitivity analyses or additional analyses | 14,16 | Results, paragraphs 8,12 |
|  | c) | Report any assessment of direction of causal relationship (e.g., bidirectional MR) | 14 | Results, paragraph 8 |
|  | d) | When relevant, report and compare with estimates from non-MR analyses | 16-18 | Discussion |
|  | e) | Consider additional plots to visualize results (e.g., leave-one-out analyses) | N/A |  |
|  | **DISCUSSION** |  |  |  |
| 14 | **Key results** | Summarize key results with reference to study objectives | 16,19 | Discussion, paragraphs 1 & 7 |
| 15 | **Limitations** | Discuss limitations of the study, taking into account the validity of the IV assumptions, other sources of potential bias, and imprecision. Discuss both direction and magnitude of any potential bias and any efforts to address them | 18,19 | Discussion, paragraph 6 |
| 16 | **Interpretation** |  |  |  |
|  | a) | Meaning: Give a cautious overall interpretation of results in the context of their limitations and in comparison with other studies | 16-18 |  |
|  | b) | Mechanism: Discuss underlying biological mechanisms that could drive a potential causal relationship between the investigated exposure and the outcome, and whether the gene-environment equivalence assumption is reasonable. Use causal language carefully, clarifying that IV estimates may provide causal effects only under certain assumptions | 16-18 |  |
|  | c) | Clinical relevance: Discuss whether the results have clinical or public policy relevance, and to what extent they inform effect sizes of possible interventions |  |  |
| 17 | **Generalizability** | Discuss the generalizability of the study results (a) to other populations, (b) across other exposure periods/timings, and (c) across other levels of exposure | 19 | Methods, paragraphs 5,6. Discussion, paragraph 6 |
|  | **OTHER INFORMATION** |  |  |  |
| 18 | **Funding** | Describe sources of funding and the role of funders in the present study and, if applicable, sources of funding for the databases and original study or studies on which the present study is based | 21-22 |  |
| 19 | **Data and data sharing** | Provide the data used to perform all analyses or report where and how the data can be accessed, and reference these sources in the article. Provide the statistical code needed to reproduce the results in the article, or report whether the code is publicly accessible and if so, where | 19 |  |
| 20 | **Conflicts of Interest** | All authors should declare all potential conflicts of interest | 22 |  |

This checklist is copyrighted by the Equator Network under the Creative Commons Attribution 3.0 Unported (CC BY 3.0) license.

1.	Skrivankova VW, Richmond RC, Woolf BAR, Yarmolinsky J, Davies NM, Swanson SA, et al. Strengthening the Reporting of Observational Studies in Epidemiology using Mendelian Randomization (STROBE-MR) Statement. JAMA. 2021;under review.

2.	Skrivankova VW, Richmond RC, Woolf BAR, Davies NM, Swanson SA, VanderWeele TJ, et al. Strengthening the Reporting of Observational Studies in Epidemiology using Mendelian Randomisation (STROBE-MR): Explanation and Elaboration. BMJ. 2021;375:n2233.
